# Eurofins Covid-19 Sentinel™ Wastewater Test Provide Early Warning of a potential COVID-19 outbreak

**DOI:** 10.1101/2020.07.10.20150573

**Authors:** Alissa Carina Udi Jørgensen, Jesper Gamst, Line Visby Hansen, Ida Ingeborg Høgh Knudsen, Søren Krohn Skovgaard Jensen

## Abstract

The Eurofins Covid-19 Sentinel^™^ program was developed to monitor the evolution of the pandemic and for early detection of outbreaks. The study objective was to develop a wastewater testing method to analyze SARS-CoV-2 as an indicator of community infection rate as of resurgence of COVID-19 in well-defined sites such as production facilities, hospitals or nursing homes. Eurofins performed >700 tests on 78 unique samples from 18 sites in Denmark, France and Belgium. Ten variant test protocols were trialed. Protocol variations trialed included centrifugation, precipitation of the SARS-CoV-2 RNA, agitation prior to precipitation, cooling, and pasteurization of the samples. A method was succesfully developed and reliability was supported by stability, reproducibility, and dilution & linearity studies. Results obtained showed a direct link to number of RNA copies in the sample using a calibration curve with synthetic SARS-CoV-2. Analysis was performed on both the liquid phase and solid phase of wastewater samples, with virus RNA detected in both phases but more frequently in the liquid phase. The virus was present in a sample from a Danish community wastewater treatment plant collected on February 24, 3 days before the first COVID-19 case was officially reported in the country. The greatest concentration of virus detected corresponded to when the COVID-19 crisis was at its peak in Denmark. Based on studies carried out in a Danish hospital, the wastewater testing method is expected to be able to detect a community COVID-19 prevalence rate as low as a 0,02%-0,1% (i.e. between 2 virus shedders per 10000 persons and 1 virus shedder per 1000). The wastewater testing method was used to monitor a Danish Community after a COVID-19 outbreak and it was shown that the method can be used as a semi-quantitative method to monitor the development of an outbreak.

## 1. Introduction

Eurofins Covid-19 Sentinel^™^ tests have been developed to monitor the evolution of the pandemic and for early detection of outbreaks. The objective of this study was to develop a wastewater testing method to analyze SARS-CoV-2 as an indicator of the infection rate in a community or in well-defined sites such as production facilities and nursing homes. Increasing the frequency of human testing has globally to date been the most common approach to track the spread of SARS-CoV-2. This invasive approach can be costly, complex, time consuming, and prone to privacy management concerns. Wastewater from a defined area can be representative of an entire community, a large part of a company or a single facility. It has been established that faeces of infected persons contain virus (Hart & Halden, 2020), even when those infected don’t show disease symptoms (i.e. are either asymptomatic or pre-symptomatic). The first Italian patient with COVID-19 was diagnosed on February 21, 2020 (Indolfi & Spaccarotella, 2020) but wastewater samples collected in Italy showed that the virus was already present in December 2019 (Istituto Superiore di Sanità, 2020). Samples collected from Wastewater Treatment Plants (WWTPs) or from wastewater generated from work sites may be useful as a leading indicator of Coronavirus presence in a community or work site. Several studies have already shown that this is possible (Medema, Heijnen, Elsinga, & Italiaander, 2020) (Wurtzer, Marecha, Mouche, & Moulin, 2020) (Stoecklin, et al., 2020). If this approach is used by individual companies or facilities, frequent monitoring of the wastewater may provide a noninvasive early indication of an outbreak in a company/facility before symptoms appear, enabling protective measures and employee PCR testing included in Eurofins SAFER@WORK^™^ programs to be effectively introduced.

## 2. Development of a SARS-CoV-2 Wastewater Test Protocol

Eurofins performed >700 tests on 78 unique samples from 18 unique sites on samples collected from Wastewater Treatment Plants (WWTP) in 11 Danish cities, 2 Danish hospitals, 1 French WWTP and 4 Belgian hospitals. A test protocol was developed based on experiences from (Stoecklin, et al., 2020), (Medema, Heijnen, Elsinga, & Italiaander, 2020), (Wu, et al., 2020) and (Wurtzer, Marecha, Mouche, & Moulin, 2020). Ten variant test protocols were trialed. The protocol variations trialed included centrifugation, precipitation of the SARS-CoV-2 RNA, agitation prior to precipitation, cooling, and pasteurization of the samples.

At first, a wastewater sample, from a hospital in Denmark known to be treating patients infected with COVID-19 was collected in order to obtain a potentially positive sample. The sample was collected as a time-dependent composite sample over 24 hours following principles in ISO 5667-10:2004 from April 29^th^ to April 30^th^ 2020. Then, the ten variant precipitation protocols were trialed and evaluated for the presence of SARS-CoV-2 using the VIR*Seek* Screen kit, developed by Eurofins Technologies (Eurofins Technologies, 2020), followed by VIR*Seek* Ident kit if Coronavirus RNA was detected. Wastewater testing was carried out at Biosafety level 2 as Murine NoroVirus (MNV) (Eurofins Technologies, 2020) was used as a process control to validate that the method is able to extract the virus from the samples.

The initial protocols were inspired by first reports of SARS-CoV-2 detections in wastewater (Wu et al 2020) and previous papers describing detection of enveloped virus from wastewater (Ye et al 2016). Protocols also considered how the virus can be precipitated, and practical constraints, e.g. centrifugation force was reduced and initial volume was kept small. In all protocols, the sample was first centrifuged to separate suspended solids/sludge from the liquids (supernatant). Only the liquid phase was used for PEG-8000 precipitation of the virus. The protocols varied in agitation time, centrifugation time, centrifugation temperature and PEG-8000 content. All combinations were tested with and without inactivation of virus by pasteurization prior to precipitation. Inactivation previously had been reported to influence efficiency of PEG-8000 precipitation of enveloped virus (Ye et al 2016).

Results of the PCR analysis from the different protocols are shown in Figure 1.

**Figure 1.**
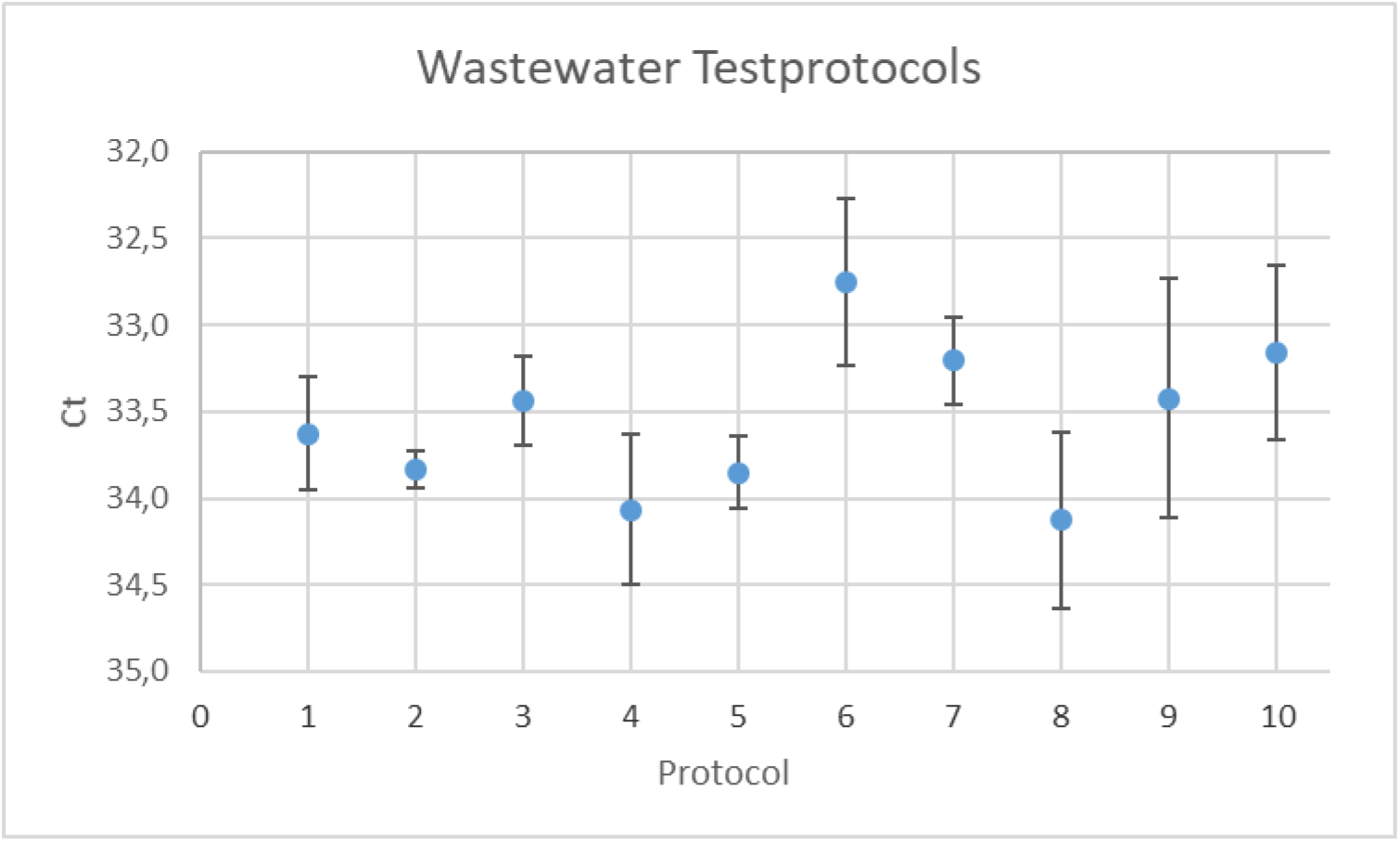
Comparison of 10 different test protocols of wastewater from a hospital in Denmark. Ct are results from PCR E-gene, all results are confirmed by RdRP gene. All protocols are tested in 4 replicates.

The best PCR signals (lowest Ct-value) for the virus pellet were obtained from Protocols 6 and 7. Because protocol 6 had fewer steps, this protocol was used for subsequent experiments. All samples from all protocols contained SARS-CoV-2. There was no clear indication that PEG-8000 content, agitation time or inactivation affected the virus detection. There was a tendency towards better detection when working at 4°C compared to 25°C.

For early stage experimental design of this study, only the liquid phase of the wastewater was to be analyzed – discarding the sludge portion during sample preparation. This assumption was based on the potential risk of interference of the PCR due to the presence of unknown chemical contaminants, cell debris, and other organic-containing matter. Neither the Eurofins primers nor the RNA extractors were previously validated for use with sludge. There is, however, no consensus in the literature for limiting testing to the liquid phase, (Wu, et al., 2020), the sludge phase (Peccia, et al., 2020) or both phases (Aarestrup, 2020). As a consequence of the lack of consensus, the testing protocol was enhanced to consider both the solid and liquid phases with consideration that one potentially could be ignored in future studies.

In Figure 2 the principles behind this protocol are shown.

**Figure 2.**
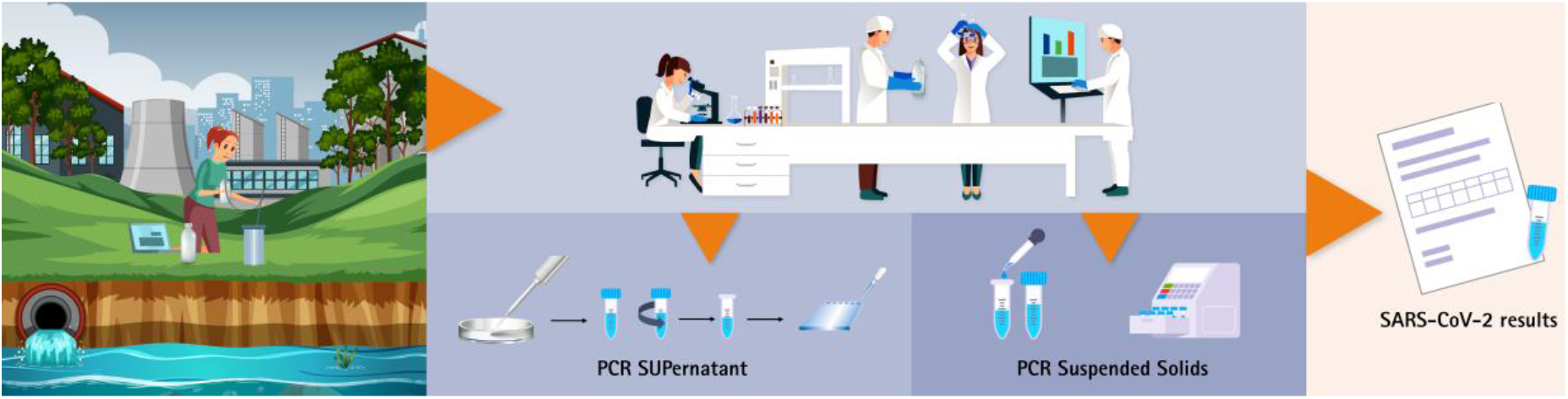
From wastewater to a SARS-CoV-2 result.

Collection of wastewater is the first step. Eurofins recommends using flow-proportional sampling or time-dependent sampling ideally following ISO 5667-10:2004. This sampling strategy generates a composite, integrated measurement of waste water and a broader picture of potential infection rate. It is strongly recommended to avoid grab-samples, which offer a very limited “snapshot” in time, unless multiple grab samples are to be analyzed within defined time periods and are expected to represent the population present on the given site.

The sample is transported directly from the sampling site to the central laboratory for the country and stored in a cold-room (4C).

Before the analysis, the samples are processed according to Protocol 6, which includes several steps to assemble two pellets – 1 from the liquid phase and 1 from the suspended solids.

The analysis was performed on both pellets referred to as the liquid phase (supernatant) and solid phase (suspended solids) using Eurofins Technologies VIR*Seek* RT-PCR tests in accordance with WHO recommendations. The test kits include a one-step real-time RT-PCR Tests for two target genes on the SARS-CoV-2 virus genome and control samples.

The signal obtained from a PCR analysis is a fluorescent intensity translated to a Ct value. The Ct value or cycle threshold value also called cycle quantification value (Cq) is the PCR cycle number at which the sample reaction curve intersects the threshold line. This line is the point at which a reaction reaches a fluorescent intensity above background levels and functions as a level of detection indicator. The Ct value is the number of cycles required to detect a discernable signal from the sample. The greater the Ct value the more cycles are required to achieve detection, which implies a lower RNA content in the specimen. This principle is represented in figure 3, which shows the amplification plots from a test performed by Eurofins.

**Figure 3.**
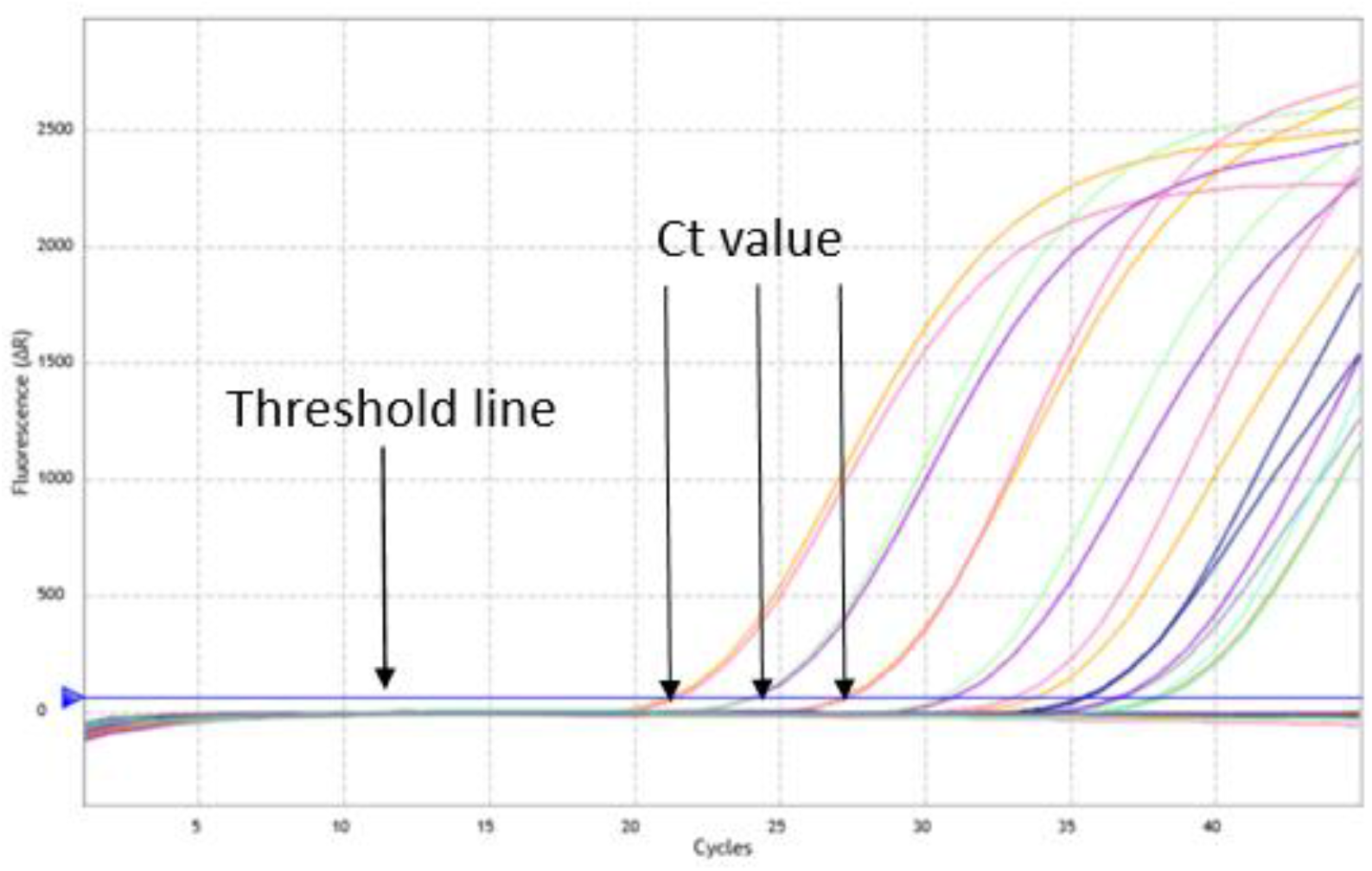
Amplification plots from different wastewater samples, illustrating thredshold line and Ct value. Each coloured line represents the fluorencent intensity from a different sample.

## 3. Validation of the protocol

Stability of SARS-CoV-2 and the reproducibility of the method was tested to confirm the validation of the protocol.

### 3.1 Sample Stability

Stability of SARS-CoV-2 RNA in the raw wastewater sample was tested by storing a wastewater sample from a Danish hospital in a cold room (4°C). Two to ten samples were analyzed on each of the experimental days during a period of 25 days. This test was conducted because a literature search did not provide consensus storage protocols for wastewater samples containing the SARS-CoV-2 virus. It is most commonly recommended to precipitate the pellet and then store the pellet at −80C. We chose to store raw wastewater samples in a cold room to test the stability of Coronavirus RNA under normal storage conditions. Figure 4 shows the Ct values obtained from the real time RT-PCR analysis performed on four different days during 25 days of storage.

**Figure 4.**
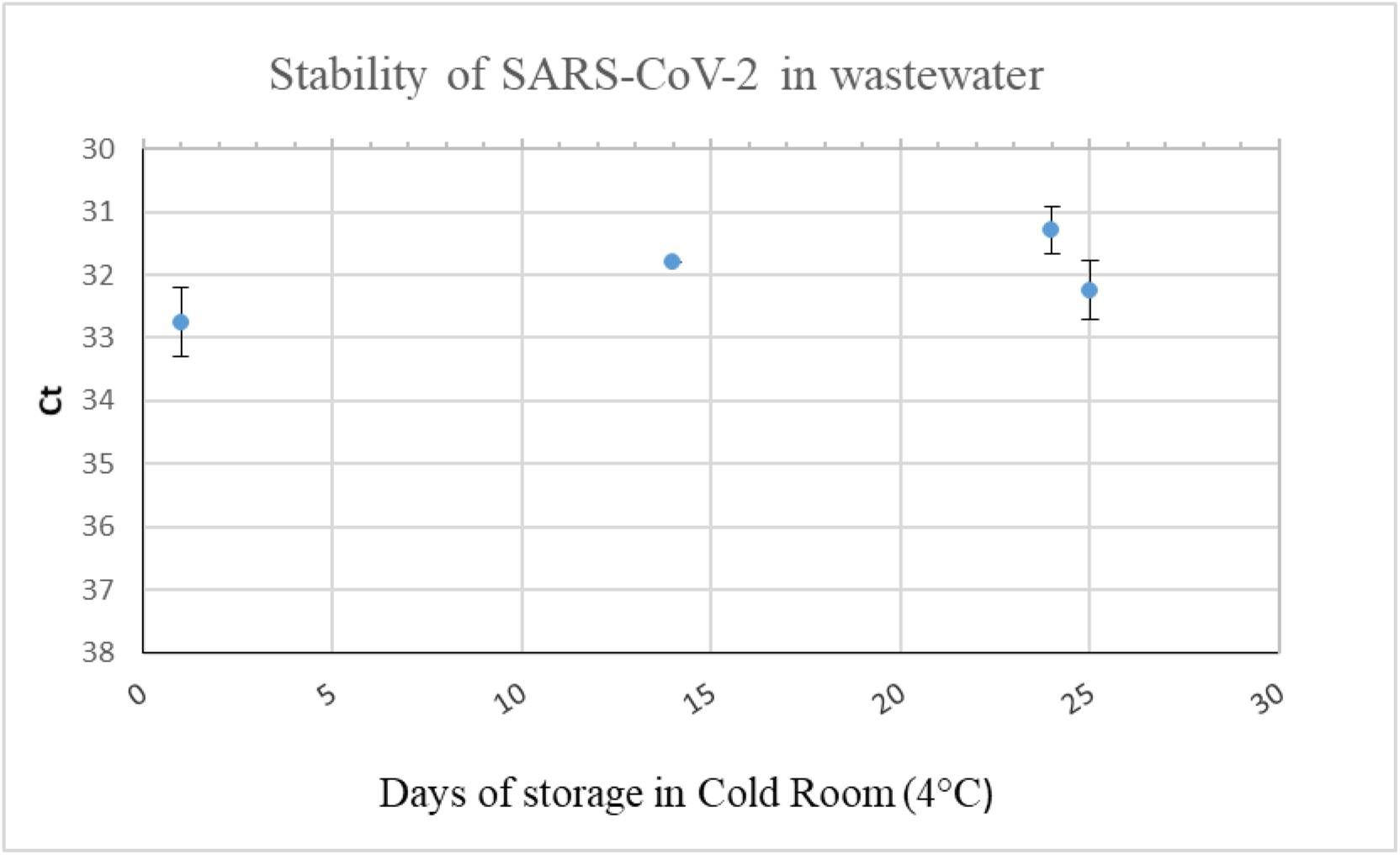
Stability of SARS-CoV-2 in wastewater stored in a cold room at 4°C for a period of 25 days.

The data supports that storing the raw wastewater samples in a cold room at 4°C does not destroy the RNA of the sample. This finding is of value where there is a need for retrospective testing of samples from a wastewater treatment plant or work site that had been previously collected and stored. The repetition experiment with 10 samples after 25 days of storage from the same container clearly shows that the method is stable within low mean standard deviations.

### 3.2 Reproducibility

Reproducibility was investigated by performing tests on both phases (liquid phase and solid phase) in 16 subsamples of a sample from a Belgian hospital. Of the total of 32 tests, RNA was detected in more than half of the subsamples, mainly in the liquid phase.

Figure 5 shows the reproducibility. Four of the tests were performed after 1 day of storage of raw wastewater sample at 4°C (square), 2 of them were performed after 6 days of storage of raw wastewater sample at 4°C (triangle) and 10 of them were performed after 13 days of storage of raw wastewater sample at 4°C (filled circle).

**Figure 3.**
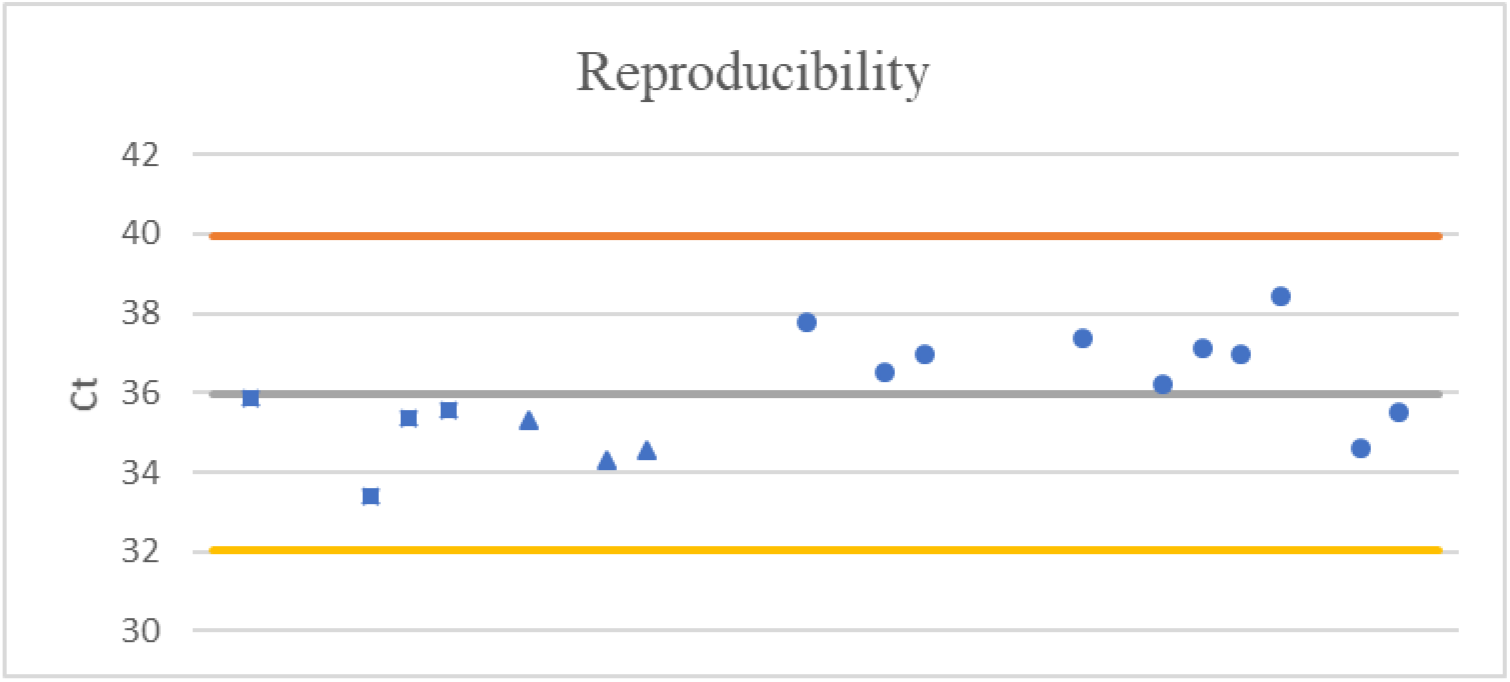
Reproducibility. Four of the tests were performed the day after sampling (square), 2 of them were performed 6 days after sampling (triangle) and 10 of them were performed 13 days after sampling (filled circle). The orange line indicates the upper limit (3*StD), the grey line indicates the average value and the yellow line indicates the lower limit (−3*StD)

The experimental data presented in Figure 5 led to an expanded uncertainty of 8%. By comparison, the uncertainty of the data presented in Figure 4 from the Danish hospital wastewater led to an expanded uncertainty of 4%. These results indicate that the uncertainty is at a low and acceptable level.

## 4. Dilution and linearity

### 4.1 Dilution

Several dilutions of a hospital (AUH) wastewater sample were performed to investigate if there was a correlation between concentration of Coronavirus RNA in the wastewater and dilution factor. Linearity as a consequence of dilution is important due to the objective to use SARS-CoV-2 in wastewater testing as an early warning indicator. We expected the wastewater from the hospital had a relatively large level of SARS-CoV-2 and thus assumed that dilution and subsequent detection was possible.

Conscious of matrix matching, a wastewater sample with no detection of RNA was used as the diluent. It was acknowledged that some matrix influence could be inevitable, but this was deemed a low risk to the experimental outcome since spiking of samples with SARS-CoV-2 was not an option during the development of the method. The tests were performed after 15, 20 and 25 days after storing the raw wastewater sample at 4°C. Figure 6 shows the dilution experiments.

**Figure 6.**
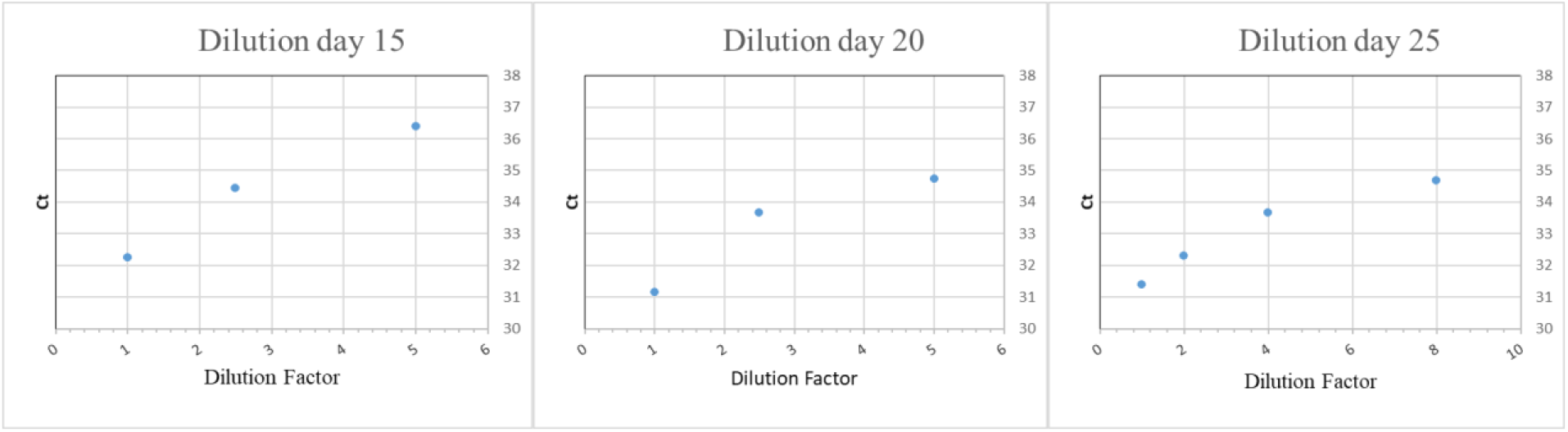
Three different dilution experiments carried out on day 15, 20 and 25, where Ct – values on the E-gene is shown as a function of dilution ratio.

Figure 6 shows a correlation between Ct values and concentration of the virus RNA (via dilution). Despite the unknown matrix effect of the diluent wastewater, the three experiments indicate that lowering the virus RNA concentration via dilution results in higher Ct values. It then follows that there are grounds to use the Ct values for quantitative analysis of virus RNA concentration, it also confirms that the method may be used to track the development (i.e. increasing viral load) of SARS-CoV-2 in a wastewater stream over time. The test work also implied that each Ct change of 1 corresponded to an approximate change in RNA concentration by a factor 2, this was especially observable on data from Day 25 on Figure 6. These dilution experiments indicate that the method can be used to track concentration variations of SARS-CoV-2 in wastewater.

### 4.2 Standard curve

Ultra pure water was spiked with standard solution of synthetic SARS-CoV-2 (Eurofins Technologies, 2020) to perform a standard curve. Figure 7 shows the standard curve obtained.

**Figure 7.**
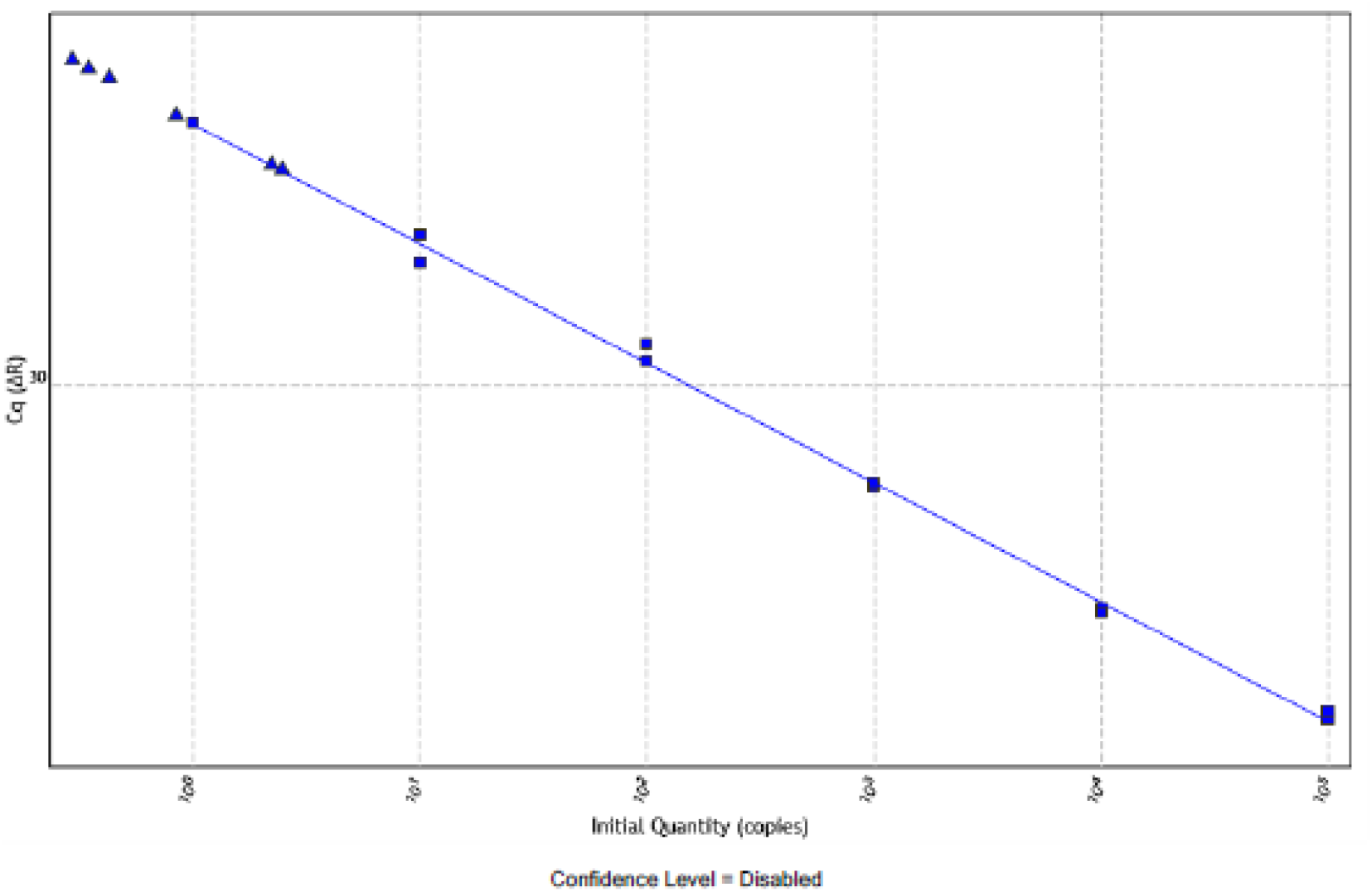
Standard curve of synthetic SARS-CoV-2.

The trend line obtained from this experiment has a slope of −3,083 and a R value of 0,997. This shows that there is a linear trend between Ct values and number of copies, making it possible to estimate the concentrator of the virus. Further tests need to be performed to verify that this trend also applies on each type of wastewater.

## 5. Supernatant and Suspended Solids

Eurofins Environment Testing Denmark stores frozen wastewater samples at −20°C for a number of months after analysis for various parameters. From this storage, 33 frozen samples collected from WWTPs during the period from February 4 till April 28 were selected. Samples from cities that recorded high COVID-19 infection rates during the crisis were targeted. In the period April 29 till June 5, 42 new samples were collected from WWTPs and hospitals.

All samples were analyzed multiple times on both pellets from the two phases using the developed test protocol. The number of samples and number of tests performed is stated in Table 2. Figure 7 illustrates the ratio of detected (both E-gene and RdRP-gene confirmation) and not detected tests.

**Table 1.** Number of samples and number of tests resulting in a detection of SARS-CoV-2 and resulting in no detection of SARS-CoV-2.

| Number of samples | Number of tests | Number of test with detection (either in solid phase, water phase or both) | Number of tests with no detection (in either of the phases) |
| --- | --- | --- | --- |
| 75 | 560 (280 on supernatant and 280 on suspended solid) | 77 | 203 |

**Table 2.** Distribution of tests, resulting in a positive detection of SARS-CoV-2, in each of the phases.

| Number of tests with detection in both solid phase and liquid phase | Number of tests with detection in solid phase only | Number of tests with detection in liquid phase only |
| --- | --- | --- |
| 21 (27%) | 15 (19%) | 41 (53%) |

SARS-CoV-2 was detected in both phases but more frequently in the supernatant (liquid). The distribution of tests, resulting in a positive detection, in the two phases is stated in Table 3 and illustrated on Figure 8.

**Figure 8.**
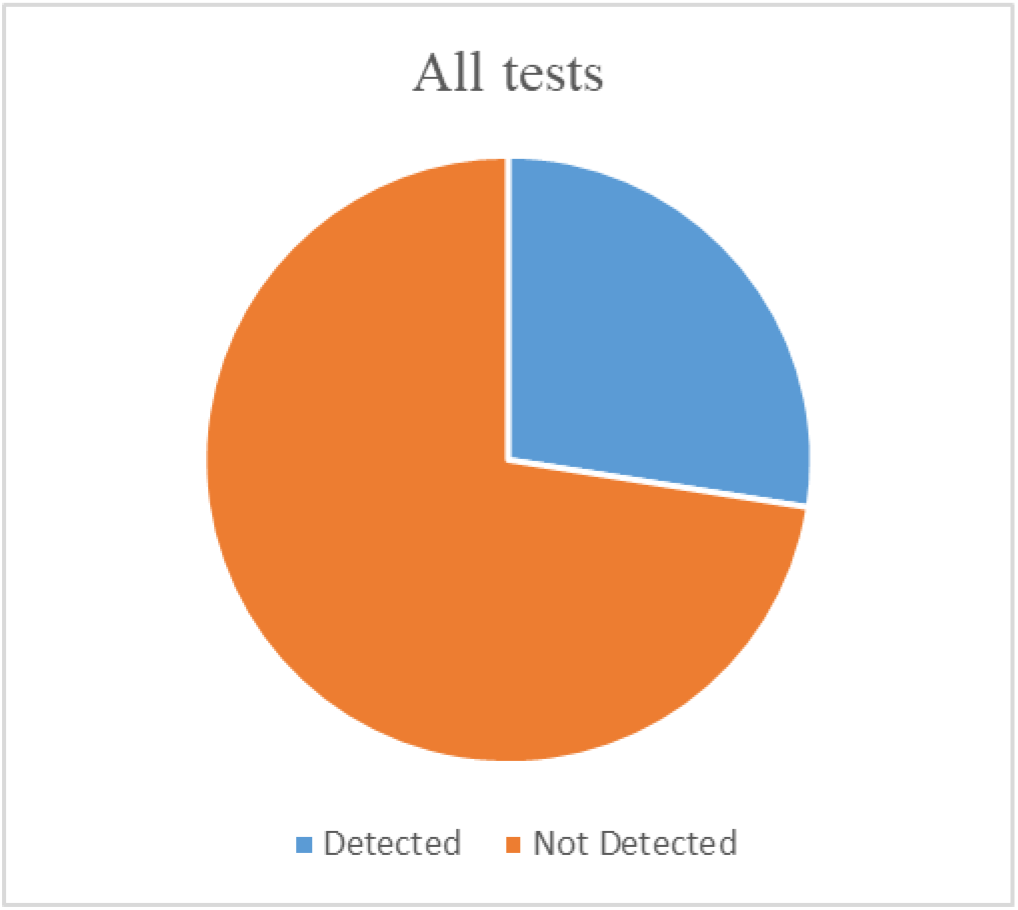
Total number of 560 tests performed on 75 unique samples. SARS-CoV-2 was detected in 77 tests and not detected in 203 tests.

Figure 9 shows that in 46% of the tests, the SARS-CoV-2 was present in the solid phase – thereby supporting the value of solid phase testing. An explanation could be the nature of the SARS-CoV-2 virus which has a lipid layer see Figure 10 (Liu, et al., 2020) and is attracted to the oil and fat found in a solid phase.

**Figure 9.**
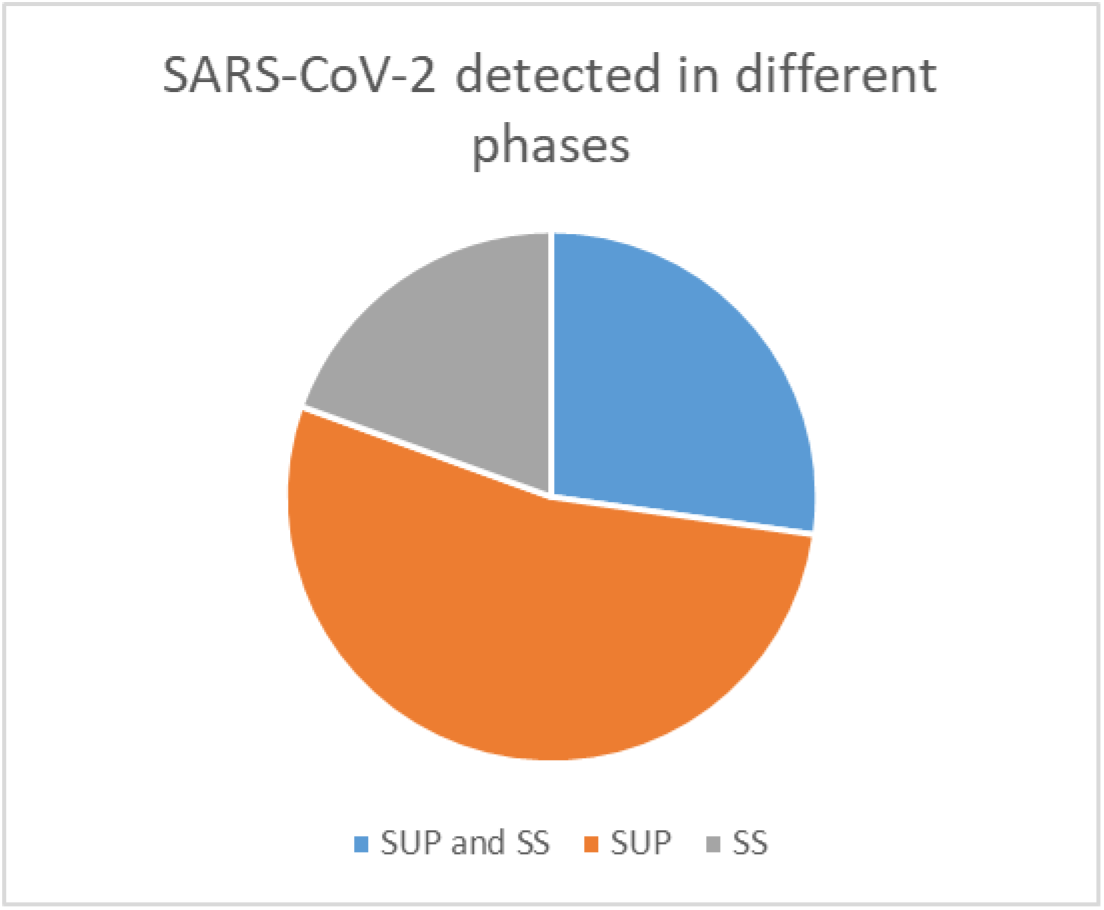
Distribution of detected RNA in the two phases. 41 in the supernatant (SUP), 15 the suspended solid (SS) and 21 in both phases.

**Figure 10.**
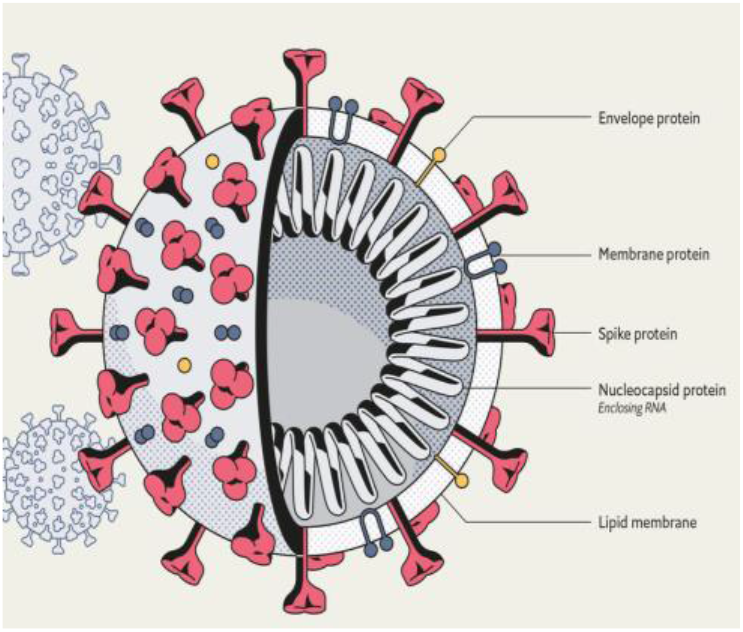
Illustration of the virus SARS-CoV-2. Mod. (Bortoletti, 2020)

Of the positive virus detections, the virus was detected in both the liquid and solid phases for 27% of samples tested and in the liquid phase for 53% of samples. In aggregate, the liquid phase was responsible for 81% of positive detections (27% + 53%). Significantly, 19% of positive detections, tested negative in the liquid phase, thereby providing a strong case for analysis of both the solid and liquid phases. Testing both phases increases the expense of testing wastewater (two PCR analyses and the double number of primers). Further method optimization should be pursued to investigate single phase analysis. This method optimization would require a technique to move the virus from the suspended solids from supernatant.

## 6. Sensitivity of the method in terms of number of infected people

The method developed to detect SARS-CoV-2 in wastewater appears robust enough to be used as a routine analytical method. For the method to be relevant to real world applications, a sensitivity analysis is required. Sensitivity analysis assumptions are provided in Figure 11.

**Figure 11.**
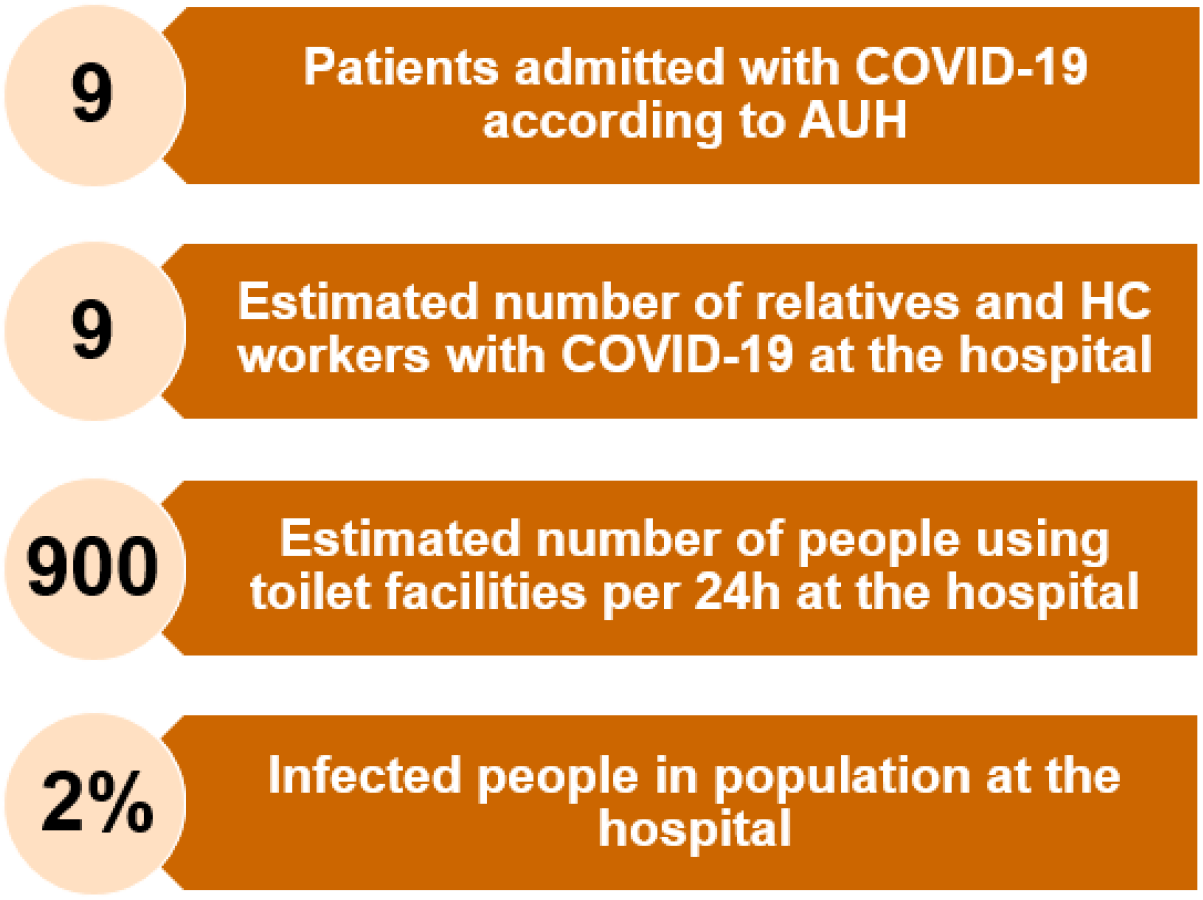
Assumptions behind estimating the sensitivity of the method.

In this assumption we used data from Aarhus University Hospital (AUH), which had nine known Covid-19 positive patients. As a best estimate to include unknown infected people, we assumed that another nine people – visitors/relatives or hospital care workers - were also infected. We assumed that all of them used the toilet during the 24-hour sampling period. In total it was estimated that 900 people used the toilet in the tested hospital - based on 800 visitors, 200 patients, and 1600 employees during the 24-hour sampling period. Apart from the nine Covid-19 patients, we acknowledge that other assumptions are premised on data provided by the hospital regarding annual numbers on coworkers and visitors and cannot be proven to be accurate. Using these assumptions, we estimate that approximately 2% (9+9 = 18 infected / 900 using toilet) of the toilet users contributing to the waste water stream were infected. The relationship between sample dilution and concentration has been established in this paper and combined with theoretical knowledge of PCR sensitivity, a method sensitivity was estimated. The threshold for the PCR analysis is approximately Ct 38 according to initial experiments. The Ct value of the hospital sample was 32. An increase in Ct value from 32 to 38 would imply that the number of virus copies in the sample would decrease by a factor of 2^6^=64 or as shown in Figure 7 close to a factor 100 (based on the slope of 3 Ct for a 10-fold change). It then follows that theoretically it should be possible to conservatively report a positive finding if even less than 1 out of 1,000 persons in the community are infected (2%/64=0,03% ∼3 pr. 10.000 or 2%/100=0,02% ∼2 pr. 10.000 persons). We acknowledge that these correlations are based on some assumptions and likely carry lower than typically accepted precision. Thus, the sensitivity of the method may be estimated to have the ability to detect in the range of 0,02-0,1% infected population (between 2 virus shedders per 10.000 persons and 1 virus shedder per 1000 persons). Thus implies that the method can be a very powerful tool for early monitoring and easily detection of COVID-19 occurrence or resurgence in a community or workplace.

## 7. Trends at several locations

SARS-Cov-2 RNA was analyzed for 38 samples sampled from 6 different WWTP, 2 Danish hospitals (AUH and RH) and 1 Belgian hospital. The samples showed several virus findings as shown in figure 11.

SARS-CoV-2 RNA was detected in wastewater from several of the tested WWTP as well as in samples collected from several of the hospital sites. Figure 12 shows data from the sites, gathered for all dates (e.g. in Solrød the wastewater is tested on samples collected on 3 different dates). The number of unique samples on each site varies from 1 to 9. Results in Figure 12 show that during the investigated period, several positive findings were detected. The results in Figure 12 indicate that the method presented in this publication is applicable for analyzing SARS-CoV-2 in wastewater. Monitoring of a Danish WWTP for 3 weeks showed that the results from the method can be translated to time stamped community observations. Figure 13 provides the trend and development of SARS-CoV-2 in the wastewater from Danish WWTP. The monitoring started when the outbreak was discovered in the community. The results show that the Ct values dropped, and thus the virus load from the community appears to have reduced. Concurrently the local authorities introduced actions to get the outbreak under control. Thus the results show that the actions introduced did have a positive impact on the virus load in the community. If continuous monitoring had been used, prior to the outbreak, the authorities could have got an early warning of the occurrence and growth of the infection cluster and the outbreak would most likely have been reduced. The results in Figure 13 also shows that the wastewater test, developed in this study, can be used as a semi-quantitative tool to monitor a development of an COVID-19 outbreak in a community.

**Figure 12.**
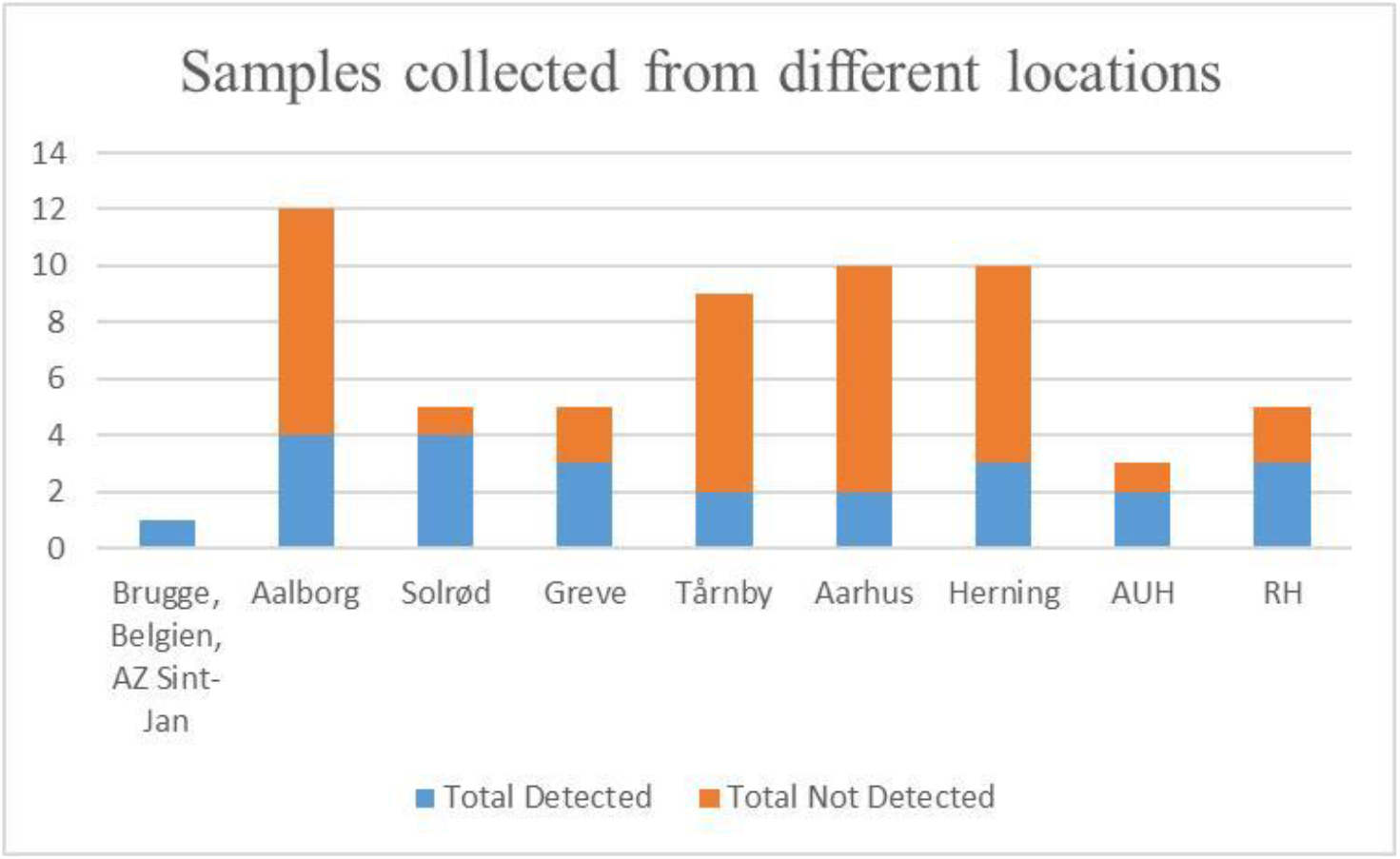
Samples, analyzed for SARS-CoV-2 RNA, from 9 different locations.

**Figure 13.**
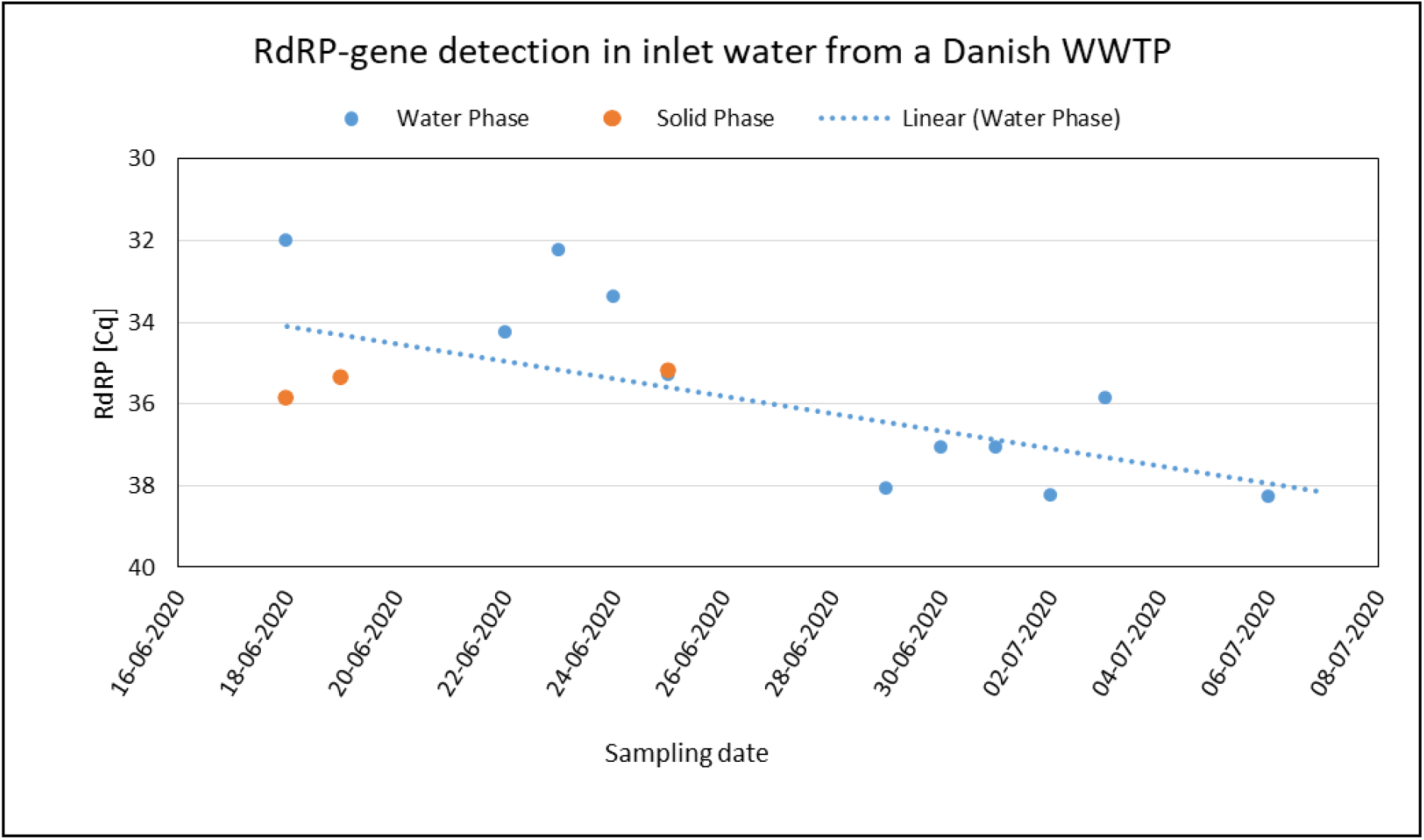
Results from monitoring a Danish WWTP for 3 weeks. The orange dots indicate results on solid phase. The blue dots indicate the results on the water phase. The dotted line shows the trend line of the water phase. Only samples with PCR signal are reported. No samples were taken on weekends.

Figure 14 shows the results for three Danish WWTPs with positive virus findings. The greatest concentration of virus is evident from the period mid to late March, corresponding to the period when the COVID-19 crisis was at its greatest peak in Denmark. Of note is the sample collected from Solrød WWTP in February 24. This sample had Coronavirus RNA three days prior to the reporting of “patient zero” from another city in Denmark on February 27. This detection further supports the value of wastewater monitoring as an early indicator of community/location infection.

**Figure 14.**
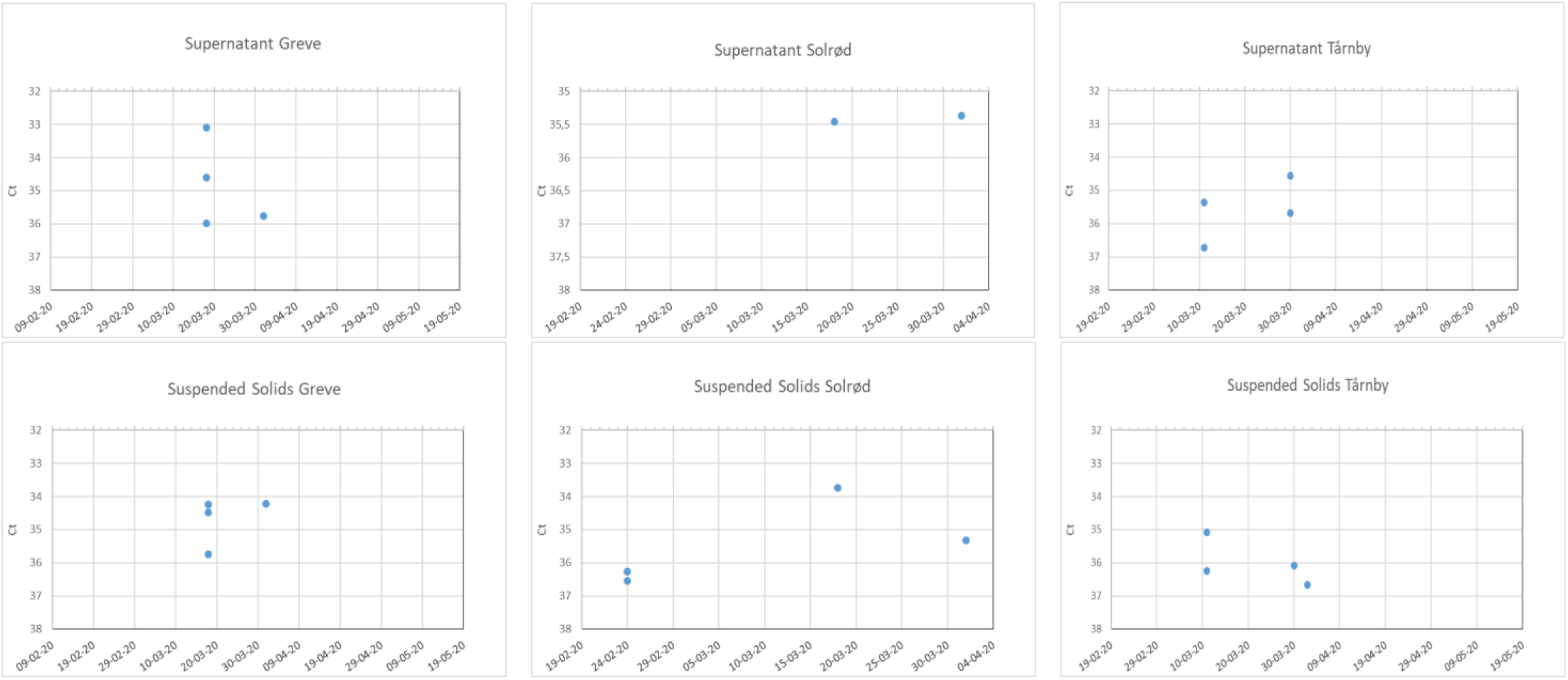
Results for 3 Danish WWTP with positive findings of virus including one before COVID-19 patient zero was detected in Denmark.

## 8. Conclusion

A method to analyze SARS-CoV-2 in wastewater samples has succesfully been developed. Method reliability is supported by stability, reproducibility, and dilution & linearity studies.

The stability of SARS-CoV-2 was tested establishing that collected samples can be stored at 4°C for 25 days without leading to any significant changes in the amount of RNA detected.

The studies support the measurement on both suspended solids and the supernatant. Developing a single phase analysis by extracting the virus from the suspended solids to the supernatant could lower the costs.

Analysis of samples collected from wastewater treatment plant inlets shows positive findings during the outbreak of the Covid-19 pandemic in Denmark. Of note is that the virus was detected in a sample 3 days before patient zero was identified in the country. The detection of SARS-CoV-2 in wastewater samples prior to the outbreak of the Covid-19 pandemic support that the method is applicable for early detection of community infection or of pandemic reocurences.

By monitoring a WWTP it was shown that the method is semi-quantitative and can be applied to follow the development of the virus load in a community during the course of a COVID-19 outbreak. Thus can be used as an early indicator of the effectiveness of public health measures to contain the virus.

The estimated sensitivity of the method is approximately 0,02%-0,1% in a community that contains infected individuals (2 positive persons in 10.000 to 1 person in 1.000). This also strongly supports the applicability of this tool as a sensitive and cost effective early indicator of infection pressence and rate.

## Data Availability

Data not available

## Acknowledgement

The authors would like to thank many colleagues in Eurofins for their effort, and support of this study. John Cosgrove Eurofins Eaton Analytical, Svend Aage Linde Eurofins Food Testing Denmark, Yvoine Remy-McCort Eurofins Environment Testing Europe, Brian Williams Eurofins Environment Testing USA, Shay Xie, Eurofins Environment Testing Australia, Saghar Mothlag, Eurofins Genescan Technologies, Arta Doci Eurofins Test America, Jens Kroll Eurofins Food Testing Germany, Yann Le Houedec Eurofins Environment Testing France, Frederic Dupont Eurofins Environment Testing Europe, Douglas Marshall Eurofins Food Testing USA, Frederic Bois Eurofins Food and Environment Testing, Gilles Martin Eurofins Scientific.

